# Deep Learning for pericardial fat extraction and evaluation on a population study

**DOI:** 10.1101/2020.01.30.20019109

**Authors:** Gianmarco Santini, Daniele Della Latta, Alessio Vatti, Andrea Ripoli, Sara Chiappino, Valeria Piagneri, Dante Chiappino, Nicola Martini

## Abstract

**Objectives:** The pericardial fat represents a powerful (promising) index which has been seen to correlate with several cardiovascular events. We propose a novel approach to automatically measure it from CT scans and we seek to explore how it is distributed in a study population.

**Methods:** We studied a population of 1528 patients where 47 (3%) showed a cardiac event. The fat segmentation model was based on a deep neural network to identify the heart and from it, the pericardial fat was extract by threshold. Statistical analysis was finally computed to stratify the population according the quantity of pericardial fat.

**Results:** The high segmentation quality was reported with a Dice index (92.5%) and a Pearson coefficient (0.990, p<0.001). Notably, normalized pericardial fat volume was significantly higher in patients with cardiac event (73.46±30.84 vs 60.06±25.38 cm^3^/m^2^, p=0.005) with a AUC of 63,44% for discrimination of cardiac event.

**Conclusions:** The proposed approach reached accurate and fast performance for the evaluation of the pericardial fat making it reliable for a population analysis.

## 1. Introduction

Pericardial adipose tissue is a metabolically and immunologically active tissue that surrounds the coronary arteries and shares with them the same innervation and blood flow. Consequently many authors seem to agree that pericardial fat can be the connection between metabolic dysregulation and vascular wall inflammation [1-4].

To date epicardial fat volume has been correlated with cardiovascular risk factors, coronary calcifications and significative coronary stenosis, and other major acute cardiac events, defined as cardiac death, non fatal myocardial infarction, and unstable angina pectoris [5-9]. It has recently been demonstrated how higher indexed volumes of epicardial tissue were correlated with high risk coronary plaques, independently of coronary calcifications (revealed as Calcium Score (CAS) levels), the degree of stenosis and classical risk factors [10]. The crucial role of inflammation in atherogenesis and plaque instability is well known. In a very recent work, Antonopoulos suggests that coronary Calcium Score, which is the only biomarker deriving from non-invasive imaging that has a predictive value in preventing coronary event, describes structural alterations of coronary wall that are irreversible and not modifiable [11]. This underlines the need to have a marker of vascular inflammation, useful for the early stratification of the risk of events. The biologic properties of adipose tissue are largely dependent on the degree of adipocytes differentiation, that is suppressed by inflammation [12]. Based on this, Antonopoulos has proven that the average attenuation level of the epicardial adipose tissue, as measured from reconstructed CT, was inversely related to the expression of adipogenic genes and to adipocyte size, identifying an imaging marker of early vascular inflammation [11].

Because of the high time required for a manual segmentation, the lack of dedicated software widely available in clinical sites, and the intra and inter observer variabilities, the pericardial fat quantification does not represent a trivial task and its measurement is not routinely performed in clinical practice.

A series of approaches have been proposed to address this problem, starting from CT acquisitions, trying to achieve an automatic segmentation that could be fast and accurate at the same time. For instance, Shahzad et al. developed an automatic multi-atlas based method to segment the pericardium and subsequently evaluate the pericardial fat from non-contrast enhanced cardiac CT scans [13]. In this work some limitations were probably related to the necessity of using CT scans with same field of view of the reference contrast-enhanced CT (CECT) scans, and to the presence of segmentation leaking as consequence of particular heart shapes. Moreover, no information about the processing time were provided. Ding et al. adopted a similar atlas approach to identify the pericardium through a series of atlas transformations combined with an active contour approach. In this case the high segmentation performance was reported, with also a fast processing time [14].

More recently E.O. Rodrigues et al. employed a machine learning strategy, based on regression algorithms to estimate the epicardial fat quantity, starting from the mediastinal adipose tissue and vice versa. Despite the high performance achieved by the algorithm, the processing time for a single patient remains high (0.9 hours) [15].

However, it is worthy to note the difficulty in visualizing the pericardium, particularly in thin patients. Therefore it is easier to measure pericardial fat rather than epicardial fat.

In this paper we adopt a strategy whereby an automatic segmentation of the pericardial fat, from only non-contrast enhanced CT scans, is performed using the latest technology of the Convolutional Neural Networks (CNNs) in order to operate a fast retrospective quantification of the aforementioned adipose tissue, as depicted in Figure 1.

**Figure 1.**
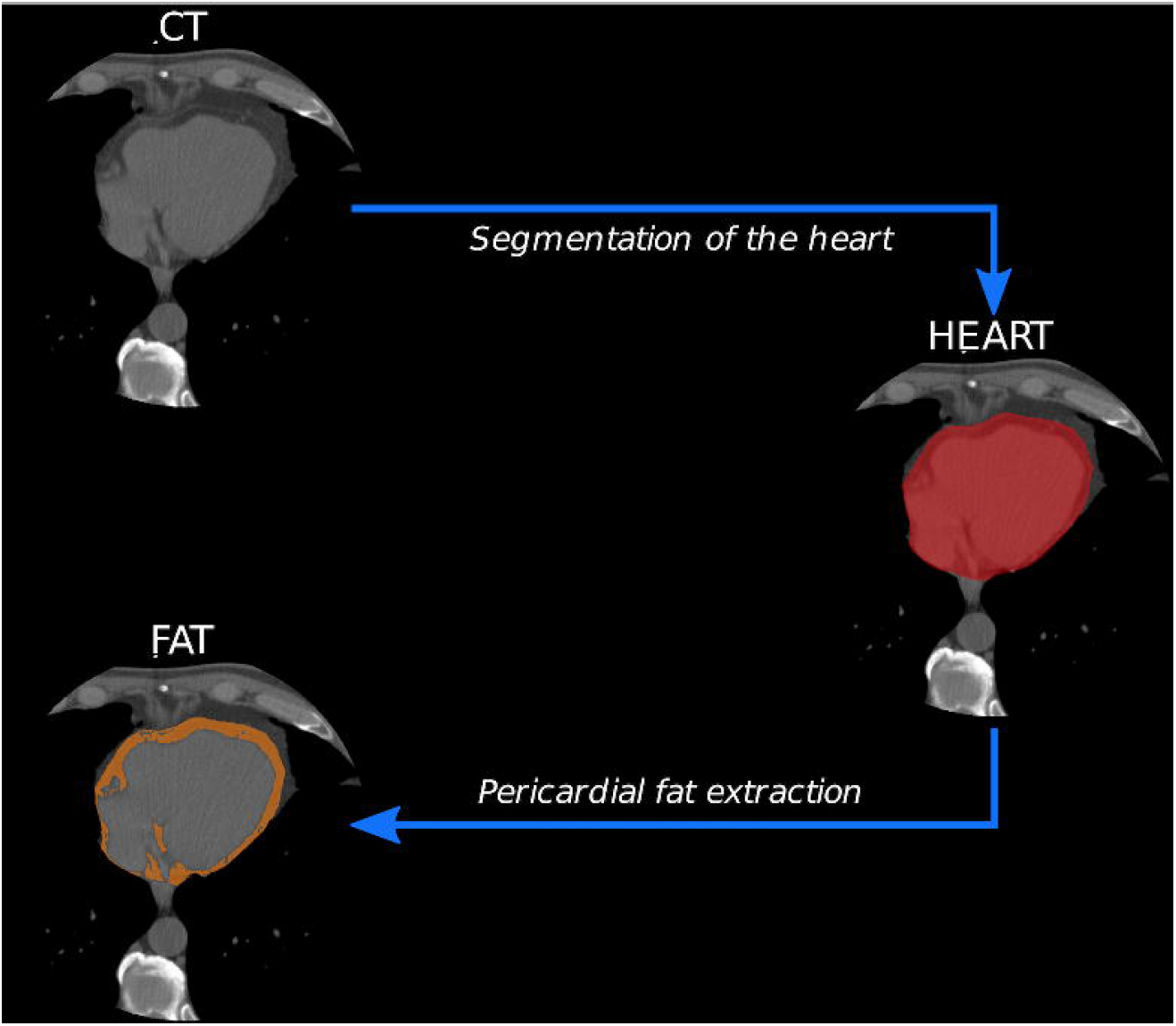
Schematic workflow of the proposed method for the extraction of the pericardial fat.

Accordingly we want to investigate how the pericardial fat measurement is distributed in a study population, where a part of it showed cardiovascular events, and prove the validity of the proposed segmentation approach as suitable tool for a clinical study.

## 2. Materials and Methods

### 2.1. Study population

The Internal Review Board and the Research Ethics Committee approved this study (registered number 2008/2514). All the participants will receive a spoken description of the aim of the study, and the choice to participate will be free and voluntary. Participants’ enrolment will occur after signing the written informed consent, including the terms of confidentiality. The results will be presented at national and international conferences andpublished in peer-reviewed journals and social media.

We collected data from a screening program (MHELP [16]) for the evaluation of cardiovascular risks in a generic population. Starting from 2010, 1528 consecutive subjects (669 males, 47.78 %) varying from 40 to 75 years of age, were enrolled. Among them only 47 patients (3.07%) showed a cardiac event. We refer to an event as the incidence of cardiovascular death or urgent revascularization for unstable angina.

All the subject enrolled, underwent a cardiac CT acquisition for the quantification of the coronary calcium score and a bio humoral exam of blood [17]. The ECG triggered CT scans were acquired without medium of contrast, according to the standard acquisition protocol used for the detection of coronary calcium, during the telediastolic cardiac phase. A tube voltage of 120 KVp and a modulated tube current of 50-500 mA were used, with a slice thickness fixed to 3.0 mm for all the acquisitions, while the axial resolution and the image filed of view (FOV) were free to vary as consequence of the patient’s heart size. In the population study, the acquisitions were collected from a 64 or 320 multi slice CT manufacture (Aquilion, Canon Inc, Yokohama Japan).

### 2.2. Physician analysis

The cardiac reference annotations were created with a semi-automatic FDA/CE approved software (Aquarius iNtuition, Terarecon Inc., Foster City US), placing point along the heart edge in the central axial slice and generating a closed line, interpolated with a spline function of order 3. The extracted contour was automatically replicated on the other axial slices to include the heart in the delimited region of interest (ROI). The shape of the final contours could be finally modified translating the ROI or creating new points to include heart portions excluded by the segmentation. The pericardial fat ground truth was finally represented by all the pixels contained in the cardiac ROI, with a Hounsfield Unit (HU) value between −190 and −30 HU [7,18].

The segmentation references were provided by two expert radiologists, with over ten years of experience each in CT imaging without contrast medium. The overall time taken to perform the whole operation ranges from five to ten minutes, depending also on the heart shape regularity.

### 2.3. Deep Learning analysis

#### 2.3.1. Automatic fat segmentation

The heart segmentation was performed using a Convolutional Neural Network designed with the standard encoder-decoder structure of a U-Net [19]. The complete architecture was summarized in Figure 2, while its outcomes in Figure 3(a-d). The encoding path comprised 4 repetitions of couples of convolutional layers followed by a 2×2 max pooling. The pooling layers considerably allowed to decrease the computational burden of the network, with a reduction of the number of weights.

**Figure 2.**
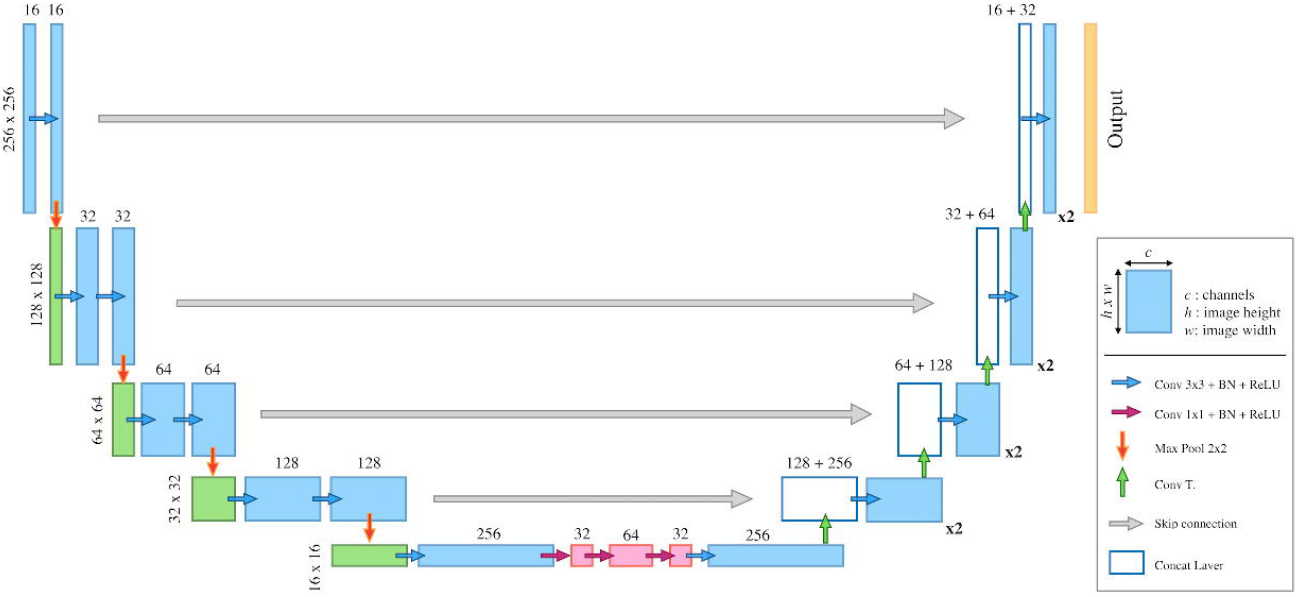
Model architecture of the designed U-Net for the heart segmentation.

**Figure 3.**
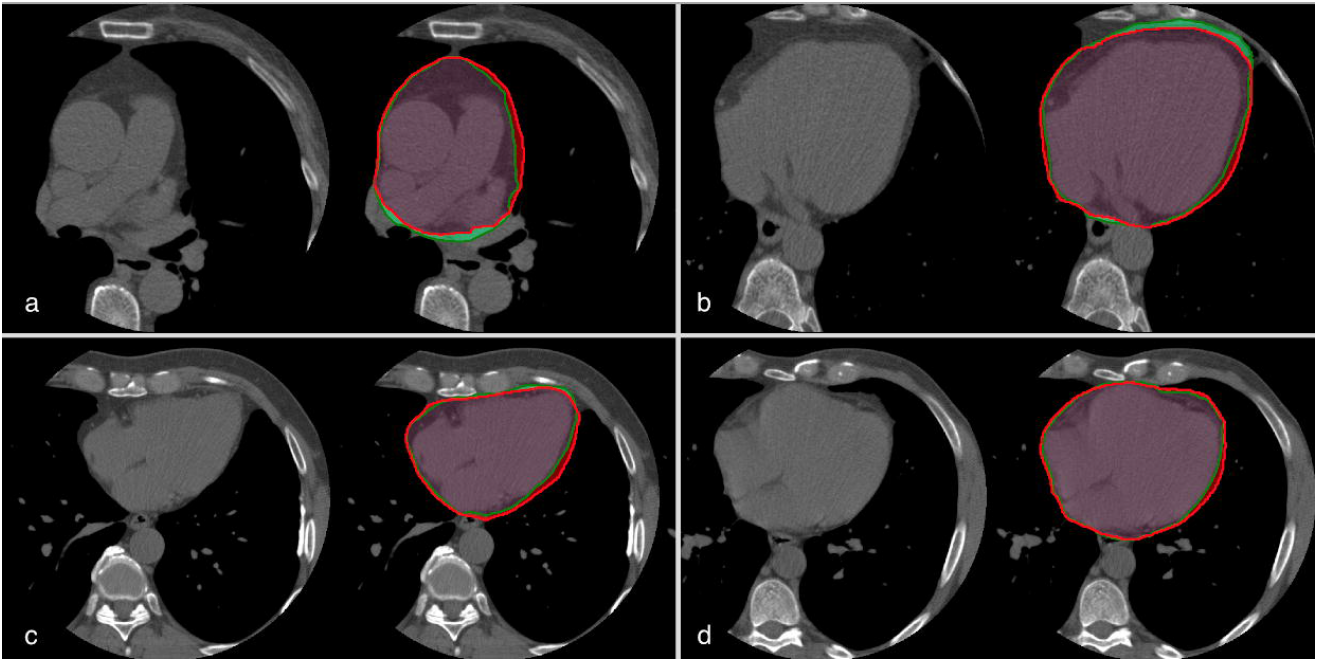
Heart segmentation examples. For each pair of images (a-d) the CT scan is shown (left) together with the ground truth heart mask (green) and the model prediction (red); the overlapping region between the two segmentation is also highlighted (violet).

For each convolution a 3×3 kernel size was initialized with a Gaussian distribution and a batch normalization was applied before every ReLU activation function to make the training process faster and less sensitive to weights initialization and learning rate setting [20, 21]. On the opposite side, the expanding path was symmetrically built using transposed convolutions with stride 2 for the up-sampling and 2 convolutional layers after each enlargement, as in the first part. While the contracting path operated a progressive reduction of the image, the expanding ones reconstructed the image original dimensions with higher level features that help to explain the image inner information. “Skip connections” were finally used to concatenate feature maps from the earlier layers with the lower resolution deeper layers, gaining good localization and use of context.

Compared to the classical U-Net architecture, where no fully connected layers were used, in our solution three 1×1 convolutional layers were added in the central part of the network in order to create an equivalent of the multi-layer perceptron and operate a dimension reduction [22]. The addition of 1×1 convolution in the architecture led to a considerable reduction of the trainable parameters in the network middle part, making there irrelevant the use of a dropout layer there, as in the original work [19]. They also allowed to create a deeper model, adding more non-linearity.

The problem of class imbalance was addressed using a weighted loss function. The weights were chosen according to the training labels occurrences normalized by the total number of pixels.

We considered the pericardial fat as the sum of both epicardial and pericardial fat [7,18]. Its extraction was realized by applying a threshold ranging from −190 to −30 HU on the heart portion segmented by the CNN (Figure 4(a-d)). The total count of pixels not excluded by the thresholding operation and multiplied by the image voxel size, corresponded to the final amount of pericardial fat.

**Figure 4.**
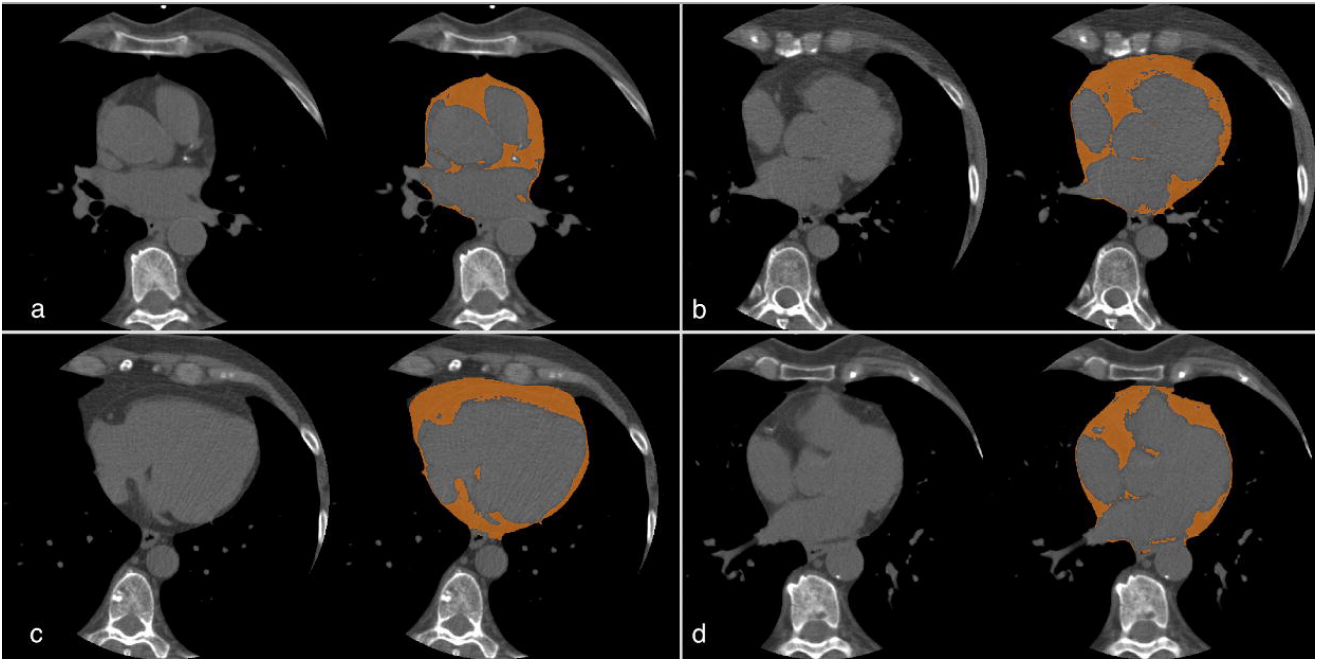
Pericardial fat extraction of randomly selected test-case slices (a-d). The orange regions show the result of thresholding the delimited heart region between −190 and −30 HU.

#### 2.3.2. Implementation details and experimental settings

We could count on a set of 119 scans from the MHELP population segmented as described in section 2.2. Among them, a random selection of 100 scans were chosen to train the deep network, designed with the intent to detect the heart in each axial image and segment the pericardial fat. The remaining 19 cases were instead used to evaluate the segmentation quality expressed by the best trained model. The two radiologists involved in the ground truth definition process, segmented different volumes of the dataset used, only once, for the training and for the test phase.

The model was implemented using TensorFlow and ran on a single NVIDIA GTX 970 for 250 epochs [23]. It was fed with two-dimensional images, taken only from the axial projection of each acquisition and reduced to 256×256 pixels with a Nearest Neighbour interpolation. The predicted masks were created with the same dimensions of the input, but to operate a direct comparison with the ground truth provided, the original dimensions (512×512) were restored with an up-sampling of a factor 2. The Adam algorithm was used as optimizer for the parameter updates, while a L2-regularized (lambda=0.001) softmax cross-entropy was chosen as loss function [24]. Finally, to further increase the training cases diversity, random rotations with a max angle of 25 degrees were applied as data augmentation.

### 2.4. Statistical Analysis

Depending on their distributions, variables extracted and considered in the retrospective analysis were reported as mean ± standard deviation, median and [25th-75th percentile] or proportion. Analogously, comparison between groups were accomplished using an unpaired t-test, a Mann-Whitney test or a chi-squared test with continuity correction, according to their distributions. The normality of continuous distributions was assessed by Kolmogorov-Smirnov test. The volume measurements were normalized by indexing to Body Surface Area (BSA). To assess the discriminative power of the pericardial fat, as predictor of cardiac event on the whole sample of patients ROC analysis was performed. A value of p less than 0.05 was considered statistically significant. All statistical computations were performed with R statistic software [25].

#### 2.4.1. Evaluation metrics

The cardiac segmentation quality was calculated on the 19 test cases using pixel-wise accuracy and Dice metric.

The agreement between the automatic estimation of the pericardial fat and the semi-automatic reference provided, was assessed by using the Pearson coefficient and the Bland-Altman with 95% confidence interval, in addition to the Mean Absolute Error (MAE) to quantify the mean volumetric difference.

Inspired by previous works the fat evaluation was performed inside of a fixed volume sub-portion [7,18]. For each subject, the considered region was extended 30 mm above and 30 mm below the central slice of the reference heart mask.

## 3. Results

### 3.1. Fat segmentation results

The presented method yielded an accuracy of 98.0% in distinguishing the pixels which belonged to the heart from the ones in the background. Moreover the Dice index computed on the test set reached a mean value of 92.5%.

The automatic measurement of the pericardial fat gained a high correlation (Pearson R: 0.990, P<0.001) (Figure 5) with the manual computed values, and a good agreement was summarized in the Bland Altman plot (Figure 6), where a bias of 0.25 cm^3^ was reported. The MAE committed in the quantification of the pericardial fat volume on all the test set cases was slightly higher than 3 cm^3^ on an average volume of 111.89 cm^3^

**Figure 5.**
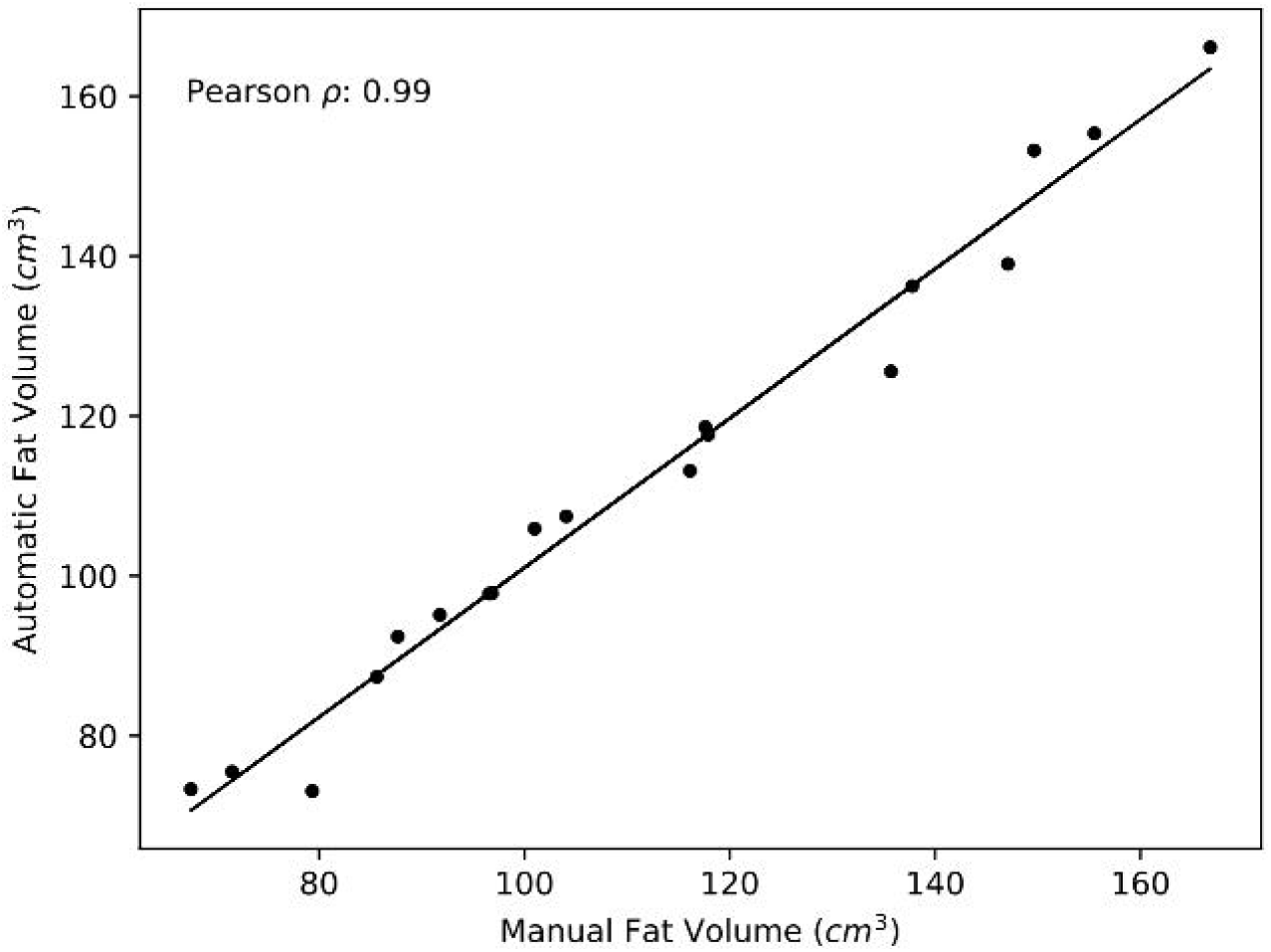
Correlation between the automatic fat estimation and the manual references provided.

**Figure 6.**
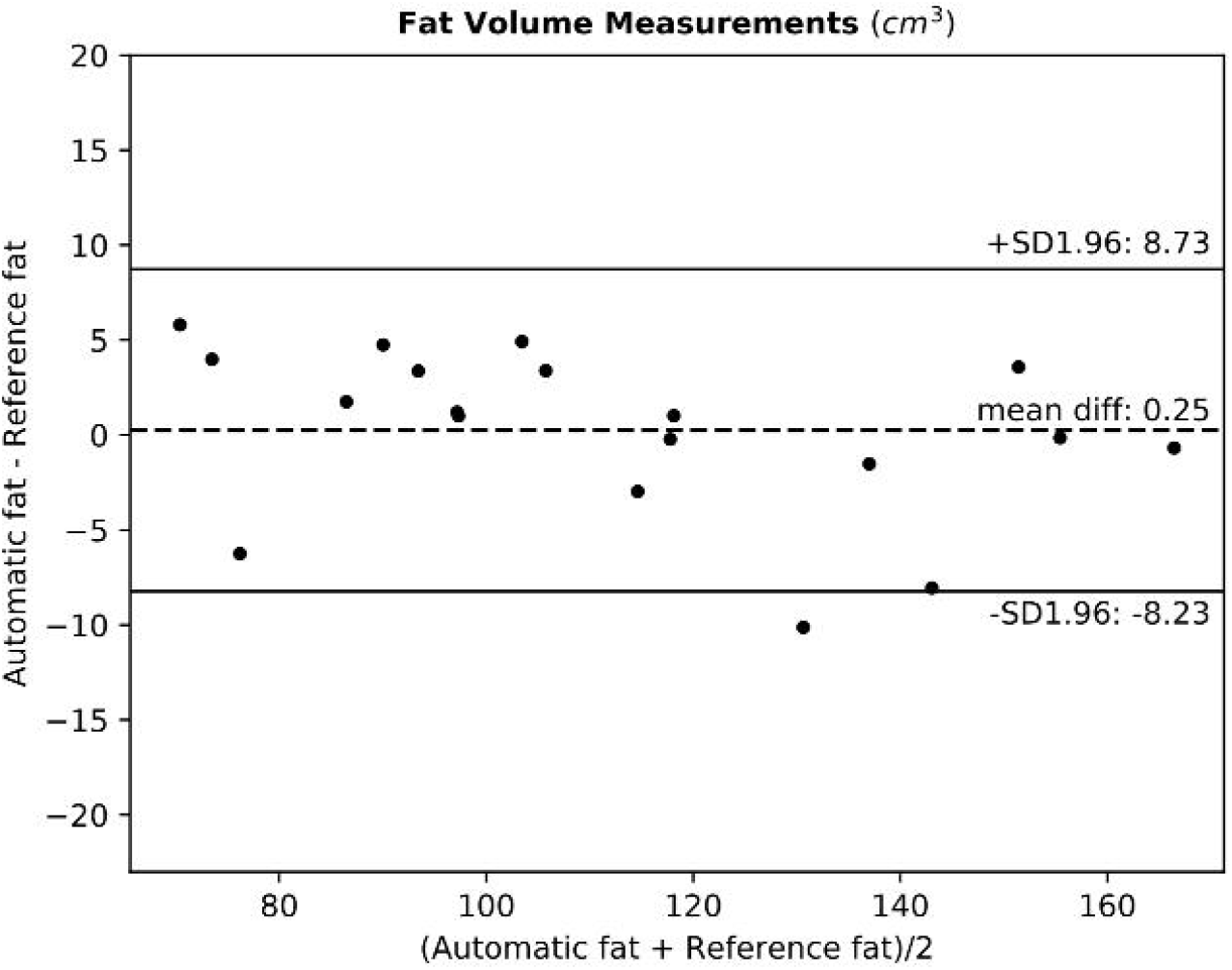
Bland-Altman plot comparing the automatically obtained fat volume with the manual references.

In terms of computational time, the trained system reached fast performance in prediction phase, with an overall segmentation and quantification time of the pericardial fat of 1.4 ± 0.3 s for each CT acquisition volume.

### 3.2. Stratification analysis

The quantification of the normalized pericardial fat was used to stratify demographics and risk factors. Results were shown in Table 1. The whole sample of patients was divided into two groups: group A (n=764), with low normalized pericardial fat, and group B (n=764), with high normalized pericardial fat. The groups were formed using the median normalized fat value (57.61 cm^3^/m^2^).

The statistical tests showed an older population characterized by higher pericardial fat volume, mainly composed by male individuals with greater weight and calcium score. Moreover, group B showed a statistically significant higher incidence of the classical coronary artery disease risk factor (hypertension, hypercholesterolemia, diabetes and hypertriglyceridemia, all with p<0.001), as well as of cardiac events (p=0.038). Results were considered in line with literature [13].

The pericardial fat was evaluated for the group of patients, who manifested a cardiac event and the group where no event was reported, giving a mean value and a standard deviation of 73.46 ± 30.51 cm^3^/m^2^ in the first case and 60.06 ± 25.38 cm^3^/m^2^ for the second case, with p=0.005.

Finally the discriminative power of pericardial fat as standalone predictor of cardiac event was investigated with ROC analysis, as reported in Figure 7.

**Figure 7:**
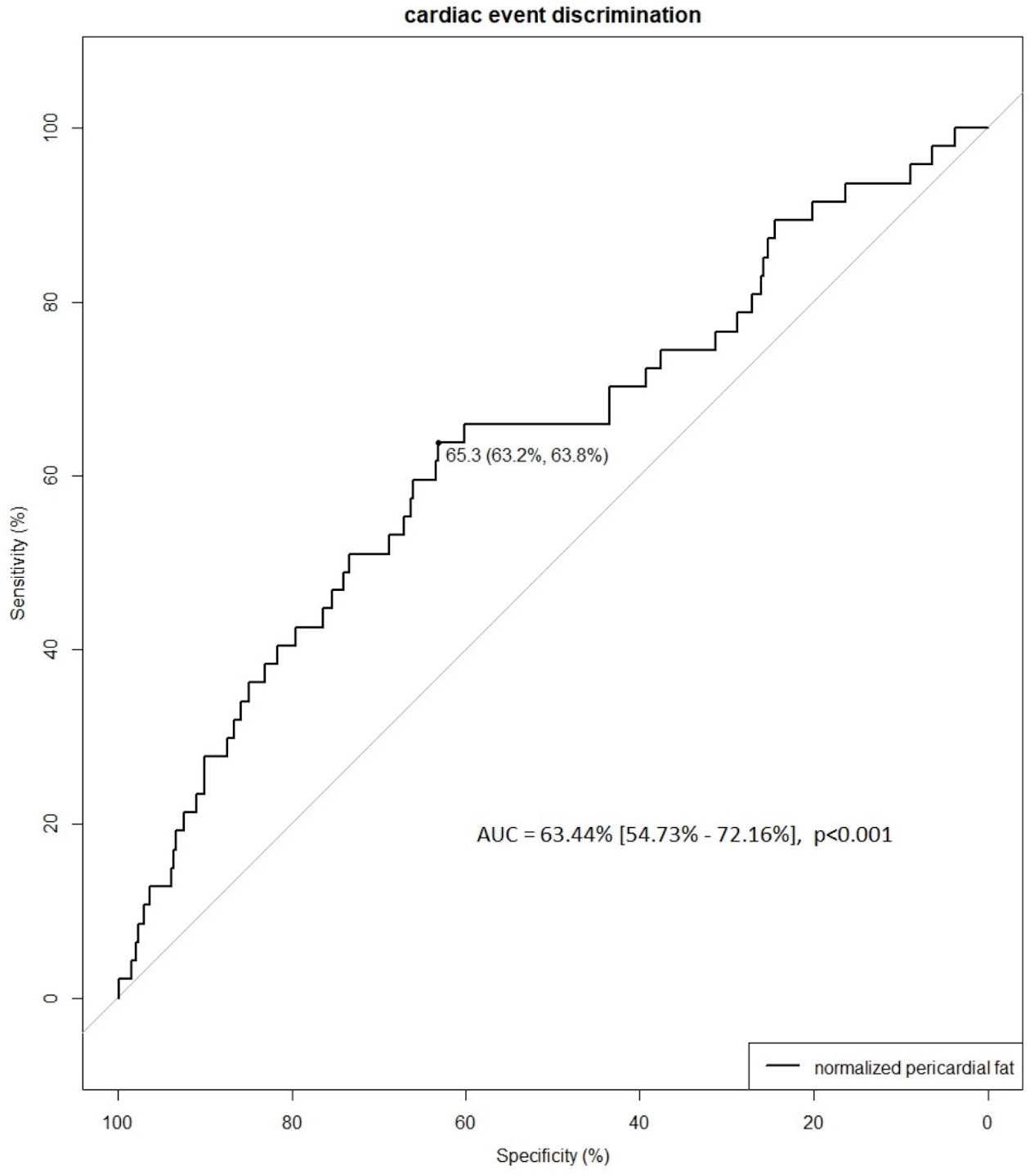
ROC curve for the discrimination of cardiac events, built with the normalized value of pericardial fat. The best threshold (65.3 cm^3^/m^2^) was computed with the Youden method.

## 4. Discussion

Many recent evidences have unveiled the association between pericardial fat and coronary artery diseases and its value in risk stratification for major cardiac events. This underlines the need to dispose of a simple and reproducible technique for pericardial fat quantification and characterization.

Here an automatic cardiac segmentation exploiting a modified U-Net was proposed as a direct solution to provide a quicker measure of the pericardial fat.

Of the whole segmentation process, the heart extraction represented the most delicate phase of the entire workflow. A wrong classification could lead to an underestimation or an overestimation of the fat volume. In this work the error committed and expressed through a MAE of ∼3 cm^3^ can be considered acceptable. The U-Net turned out to be a good choice to achieve a good heart segmentation in terms of quality and time, given the limited number of training cases. To overcome this limit, we chose to use 2D slices instead of 3D volumes as single training cases and to randomly perform on them data augmentation, reaching a considerable increase in the number and in the variability of the training samples indeed. In addition, the image variability given by the different resolutions and FOVs, the types of reconstruction filters and acquisition manufactures, represented all factors that directly contributed to further improve the U-Net capability to well generalize on the test images, avoiding overfitting incidence.

The proposed approach turn out to be not only accurate but also fast. The overall runtime in fact, comprised of the heart segmentation and the pericardial fat extraction, was significantly faster than the mean time required by an expert radiologist (1.4 sec vs 5 min).After testing the model, it was used on the entire cohort of subjects available. We considered as part of the analyzed population even the 100 cases used to train the algorithm. The fat assessment on them, did not represent a term that could significantly influence on the final results, since the 100 cases represented only the 6% of the population.

The results obtained confirmed the feasibility of the proposed approach to investigate pericardial fat distribution as consequence of the coherence revealed with the outcomes obtained in the MESA study population. In [7] the baseline characteristics for the pericardial fat returned a mean measure with a standard deviation of 79 ± 42 (cm^3^) and 100 ± 51 (cm^3^) for the “No event” and the “Event” class respectively. On our population, the values of fat extracted by using the designed approach, after removing the BSA normalization factor, were 111 ± 47 (cm^3^) and 135 ± 56 (cm^3^) for the two classes respectively. But considering that we calculated the pericardial fat on a bigger heart portion along the axial direction (18.0 cm vs 13.5 cm), the results obtained can be considered in line with the ones reported for the MESA population.

Even if computed on a different population the study also showed how the quantity of pericardial fat correlates with the cardiac events. As a matter of fact in last row of Table 1 is shown how almost twice the number of subjects reporting a cardiac event, have a greater quantity of pericardial fat than the median population. In the ROC curve also a peak value of 63.44%, for the AUC is reached. This value explain the discriminative power of the pericardial fat as standalone marker. Of course the power of predicting a cardiac event may further increase combining it with other factors.

In future works we aim to increase the number of cases to be used as learning basis for the automatic algorithm, in order to reduce the use of synthetic data produced by data augmentation and to experiment a volumetric segmentation instead of a two dimensional one. The second aspect to improve may regard the enrichment of bio-humoral marker, limited in this work to general patient information and classical coronary artery disease risk factor, and apply this approach for the local variations analysis of the average attenuation level of perivascular adipose tissue, recently identified as an imaging marker of early vascular inflammation and a vulnerable atherosclerotic plaque leading to coronary artery events.

## 5. Conclusion

In this work a new approach was presented for the extraction and the quantification of the pericardial fat by using a deep learning strategy based on convolutional neural networks. Specifically, the proposed model employs a modified U-Net architecture to extract highly meaningful feature maps and to reconstruct the semantic segmentation of the heart in basal CT scans. A threshold operation is performed to finally quantify the cardiac fat.

The method has been evaluated on a small cohort of subject providing a high agreement with the human ground truth in a shorter time. It also has been applied on a larger cohort of subject, showing to return reliable results in a population study, coherent with literature.

## Data Availability

The dataset analysed during the current study is not publicly available but is available from the corresponding author on reasonable request.

## Acknowledgements

This research did not receive any specific grant from funding agencies in the public, commercial, or not-for-profit sectors.

